# Studying the pathophysiology of coronavirus disease 2019 – a protocol for the Berlin prospective COVID-19 patient cohort (Pa-COVID-19)

**DOI:** 10.1101/2020.05.06.20092833

**Authors:** Florian Kurth, Maria Roennefarth, Charlotte Thibeault, Victor M. Corman, Holger Müller-Redetzky, Mirja Mittermaier, Christoph Ruwwe-Glösenkamp, Alexander Krannich, Sein Schmidt, Lucie Kretzler, Chantip Dang-Heine, Matthias Rose, Michael Hummel, Andreas Hocke, Ralf H. Hübner, Marcus A. Mall, Jobst Röhmel, Ulf Landmesser, Burkert Pieske, Samuel Knauss, Matthias Endres, Joachim Spranger, Frank P. Mockenhaupt, Frank Tacke, Sascha Treskatsch, Stefan Angermair, Britta Siegmund, Claudia Spies, Steffen Weber-Carstens, Kai-Uwe Eckardt, Alexander Uhrig, Thomas Zoller, Christian Drosten, Norbert Suttorp, Martin Witzenrath, Stefan Hippenstiel, Christof von Kalle, Leif Erik Sander

**Affiliations:** Department of Infectious Diseases and Respiratory Medicine, Charité – Universitätsmedizin Berlin, corporate member of Freie Universität Berlin, Humboldt-Universität zu Berlin, and Berlin Institute of Health, Berlin, Germany; Department of Tropical Medicine, Bernhard Nocht Institute for Tropical Medicine & I. Department of Medicine, University Medical Center Hamburg-Eppendorf, Hamburg, Germany; Clinical Study Center (CSC), Berlin Institute of Health, and Charité – Universitätsmedizin Berlin, corporate member of Freie Universität Berlin, Humboldt-Universität zu Berlin, Berlin, Germany; Institute of Virology, Charité – Universitätsmedizin Berlin, corporate member of Freie Universität Berlin, Humboldt-Universität zu Berlin, and Berlin Institute of Health, Berlin, Germany; Department of Psychosomatic Medicine, Charité - Universitätsmedizin Berlin, corporate member of Freie Universität Berlin, Humboldt-Universität zu Berlin, and Berlin Institute of Health, Berlin, Germany; Central Biobank Charité (ZeBanC), Institute of Pathology, Charité Universitätsmedizin Berlin, corporate member of Freie Universität Berlin, Humboldt-Universität zu Berlin, and Berlin Institute of Health, Berlin, Germany; Department of Pediatric Pulmonology, Immunology and Critical Care Medicine, Charité - Universitätsmedizin Berlin, corporate member of Freie Universität Berlin, Humboldt-Universität zu Berlin, and Berlin Institute of Health, Berlin, Germany; Department of Cardiology, Charité - Universitätsmedizin Berlin, corporate member of Freie Universität Berlin, Humboldt-Universität zu Berlin, and Berlin Institute of Health, Berlin, Germany; Medical Department, Division of Cardiology, Campus Virchow-Klinikum, Charité - Universitätsmedizin Berlin, corporate member of Freie Universität Berlin, Humboldt-Universität zu Berlin, and Berlin Institute of Health, Berlin, Germany; Department of Neurology with Experimental Neurology and Center for Stroke Research Berlin, Charité - Universitätsmedizin Berlin, corporate member of Freie Universität Berlin, Humboldt-Universität zu Berlin, and Berlin Institute of Health, Berlin, Germany; Department of Endocrinology and Metabolism, Charité - Universitätsmedizin Berlin, corporate member of Freie Universität Berlin, Humboldt-Universität zu Berlin, and Berlin Institute of Health, Berlin, Germany; Institute of Tropical Medicine and International Health Berlin, Charité - Universitätsmedizin Berlin, corporate member of Freie Universität Berlin, Humboldt-Universität zu Berlin, and Berlin Institute of Health, Berlin, Germany; Department of Hepatology and Gastroenterology, Charité - Universitätsmedizin Berlin, corporate member of Freie Universität Berlin, Humboldt-Universität zu Berlin, and Berlin Institute of Health, Berlin, Germany; Department of Anaesthesiology and Intensive Care Medicine, Charite Campus Benjamin Franklin, Charité - Universitätsmedizin Berlin, corporate member of Freie Universität Berlin, Humboldt-Universität zu Berlin, and Berlin Institute of Health, Berlin, Germany; Medical Department, Division of Gastroenterology, Infectious Diseases, Rheumatology, Charité - Universitätsmedizin Berlin, corporate member of Freie Universität Berlin, Humboldt-Universität zu Berlin, and Berlin Institute of Health, Berlin, Germany; Department of Anesthesiology and Operative Intensive Care Medicine, Charité - Universitätsmedizin Berlin, corporate member of Freie Universität Berlin, Humboldt-Universität zu Berlin, and Berlin Institute of Health, Berlin, Germany; Department of Nephrology and Internal Intensive Care Medicine, Charité - Universitätsmedizin Berlin, corporate member of Freie Universität Berlin, Humboldt-Universität zu Berlin, and Berlin Institute of Health, Berlin, Germany

**Keywords:** SARS-CoV-2, COVID-19, patient registry, infectious disease, clinical phenotyping, coronavirus

## Abstract

**Purpose:** Severe acute respiratory syndrome coronavirus 2 (SARS-CoV-2) has spread worldwide causing a global health emergency. Pa-COVID-19 aims to provide comprehensive data on clinical course, pathophysiology, immunology and outcome of COVID-19, in order to identify prognostic biomarkers, clinical scores, and therapeutic targets for improved clinical management and preventive interventions.

**Methods:** Pa-COVID-19 is a prospective observational cohort study of patients with confirmed SARS-CoV-2 infection treated at Charité - Universitätsmedizin Berlin. We collect data on epidemiology, demography, medical history, symptoms, clinical course, pathogen testing and treatment. Systematic, serial blood sampling will allow deep molecular and immunological phenotyping, transcriptomic profiling, and comprehensive biobanking. Longitudinal data and sample collection during hospitalization will be supplemented by long-term follow-up.

**Results:** Outcome measures include the WHO clinical ordinal scale on day 15 and clinical, functional and health-related quality of life assessments at discharge and during follow-up. We developed a scalable dataset to (*i*) suit national standards of care (*ii*) facilitate comprehensive data collection in medical care facilities with varying resources and (*iii*) allow for rapid implementation of interventional trials based on the standardized study design and data collection. We propose this scalable protocol as blueprint for harmonized data collection and deep phenotyping in COVID-19 in Germany.

**Conclusion:** We established a basic platform for harmonized, scalable data collection, pathophysiological analysis, and deep phenotyping of COVID-19, which enables rapid generation of evidence for improved medical care and identification of candidate therapeutic and preventive strategies. The electronic database accredited for interventional trials allows fast trial implementation for candidate therapeutic agents.

## 1. Introduction

Lower respiratory tract infections cause several millions of deaths every year and remain the most deadly communicable disease as reported by WHO (Global Health Observatory (GHO) data of the WHO, 2018). Comprehensive phenotyping of diseases caused by new infectious agents is required in order to enhance the understanding of their pathophysiology, to improve clinical management and to develop therapeutic agents. In the past 20 years alone, zoonotic viruses have caused multiple outbreaks, which all involved respiratory infections. In November 2003, Severe Acute Respiratory Syndrome (SARS) - virus caused a global disease outbreak with > 8000 infections and 774 deaths [1]. In 2012, the first infection with a new Coronavirus, the Middle-East Respiratory Syndrome Coronavirus (MERS-CoV) was recorded [2]. Despite global spread by travellers [3, 4], MERS-CoV remains largely confined to the Arabian Peninsula. In December 2019, first cases of what was later identified as infections with a novel coronavirus, termed SARS-CoV-2, emerged in Wuhan, China [5]. The local outbreak has rapidly evolved into a global health emergency (WHO Director-General’s statement, 30^th^ January 2020). SARS-CoV-2 is highly contagious by human-to-human transmission [6, 7] and can be transmitted by pre- or oligo-symptomatic individuals [8]. According to WHO, by the middle of April, there are more than 2.000.000 confirmed infections with >120.000 deaths worldwide.

Rapid generation of meaningful evidence for improved clinical management of COVID-19 requires harmonized phenotyping and comparable data collection in a well-defined patient population. Besides decade-long experience in multicentric phenotyping and clinical studies of community acquired pneumonia (CAPNETZ, PROGRESS, CAPSyS) [9, 10], Berlin has strong expertise in the areas of coronavirus biology and diagnostics [11-17], infectious diseases and highly contagious diseases, immunology of infections and vaccines, respiratory medicine and intensive care, and clinical management of ARDS [18, 19].

This unique local expertise together with the global burden of COVID-19 imply a special responsibility in assessing all available information drawn from medical care and human biomaterial within Charité as well as gathering patient data nationwide to enable fighting the outbreak of SARS-CoV-2 infection in a systematic and effective way.

*The aim of Pa-COVID-19:* Harmonized deep clinical, molecular and immunological phenotyping in patients with COVID-19 to (i) elucidate the pathophysiology of the disease (ii) identify diagnostic and prognostic scores and biomarkers for improved clinical management, (iii) identify putative therapeutic targets, (iv) produce evidence regarding short- and long-term clinical outcomes and (v) identify correlates of protective immunity. Longitudinal data- and bio-sampling as well as post-mortem analysis will enable for characterization of clinical features of distinct disease courses and identify determinants of severity and transmission.

## 2. Methods

### 2.1 Study design

Pa-COVID-19 is a prospective observational cohort study. All patients diagnosed with COVID-19 at Charité - Universitätsmedizin Berlin are eligible for inclusion. The protocol was developed in accordance with and extends upon the standardized protocol for the rapid, coordinated clinical investigation of severe acute infections by pathogens of public health interest published by the International Severe Acute Respiratory and Emerging Infection Consortium (ISARIC) [20].

We aimed to establish a study protocol that can serve as blueprint and central platform for harmonized deep clinical, molecular and immunological phenotyping studies in COVID-19 patients in Germany. The protocol supports data sharing efforts through adherence to international data harmonization standards. Data is collected longitudinally from patients with confirmed COVID-19 three times per week during their hospitalization and at follow-up visits. Data include epidemiological and demographic parameters, medical history and potential risk factors, documentation of standard of care procedures and clinical course, including different patterns of organ involvement, quality of care, morbidity and quality of life. Moreover, extensive serial high quality bio-sampling consisting of various sample types with deep molecular, immunological and virological phenotyping through single-cell- and bulk- multi-omics analysis is performed.

Here, we provide a comprehensive protocol for the interdisciplinary characterization of the syndrome caused by infection with SARS-CoV-2, a newly-emerged zoonotic virus and poorly characterized human pathogen. Therefore, the sample size is not prospectively determined. Recruitment of participants will depend on the emergence and spread of the disease in Berlin, Germany and on the number of patients presenting at Charité - Universitätsmedizin Berlin. With the evolving outbreak, recruitment in other study centres will be considered in order to facilitate the establishment of a comprehensive clinical and molecular database. The study has no set end date.

### 2.2 Inclusion criteria

Proven infection with SARS-CoV-2 (positive pathogen testing). Willingness to participate in the study.

### 2.3 Exclusion criteria

Refusal to participate by patient, parent or appropriate legal representative.

Any conditions that prohibit supplemental blood-sampling.

### 2.4 Data Collection

#### Harmonized scalable dataset / electronic Case Report Form (eCRF)

This protocol outlines the methodology of the Berlin prospective cohort for capturing data of COVID-19. The parameters were selected and adapted to local standards through a multidisciplinary expert review board consisting of clinical researchers at Charité, Department of Infectious Diseases and Respiratory Medicine, Institute of Virology, Departments of Anaesthesiology and Operative Intensive Care Medicine, Department of Psychosomatic Medicine, Department of Pediatrics, Departments of Cardiology, Department of Neurology and Center for Stroke Research, Department of Nephrology and Medical Intensive Care, Department of Endocrinology and Metabolism, Department of Medicine, Division of Gastroenterology, Infectious Diseases and Rheumatology, Department of Hepatology and Gastroenterology, Institute of Tropical Medicine and International Health, Central Biobank Charité (ZeBanC) and the Clinical Study Centre at the Berlin Institute of Health, based on available preliminary data and in accordance with the WHO supported case report form (CRF) proposed by the International Severe Acute Respiratory and Emerging Infection Consortium (ISARIC). Items of the ISARIC–CRF were translated to German using the standardized Functional Assessment of Chronic Illness Therapy (FACIT) translation methodology [21]. Data elements were prioritized into three categories (core data, recommended data and supplemental data elements, see eCRF, supplementary file 1) to allow adjustment of data collection to different healthcare and research resources.

In addition to medical history and history of present illness we aim for detailed longitudinal documentation of (i) clinical symptoms, (ii) vital signs and blood gases, (iii) laboratory parameters (including inflammation markers, cardiac enzymes, coagulation parameters etc.) (iv) imaging data (e.g. X-ray, echocardiography, ultrasound, CT-Scans, MRI-Scans), (v) microbiological and virological tests (vi) supplementary diagnostic data (e.g. bronchoscopy, lumbar puncture), (vi) concomitant medication and (vii) details of clinical interventions (mechanical ventilation, organ replacement therapy etc.) (Table 1). During in-patient stay, the patient-reported health status will be measured using tailored short-forms from the Patient-Reported Outcome Measurement Information System® (PROMIS), targeting for precise assessments of dyspnoea and physical function [22]. Overall outcomes will be evaluated using clinician as well as patient-reported health data, including the WHO 7 category ordinal scale [23] at study visits until day 15 and at discharge from hospital. Quality of life, health status and performance in activities of daily living will be assessed using the Barthel-Index and an adapted version of the PROMIS®-29 Profile at discharge, six weeks, three months, six months and 12 months after inclusion. In addition, lung function, blood gases, cardiac function, renal function, occurrence of Post-Intensive Care Syndrome, neuropsychiatric function, exercise capacity and cognitive function will be assessed in selected patient groups at follow-up visits [24].

**Table 1:** Study procedures: each visit during the patient’s hospital stay will register as a new visit. Patients that remain in hospital beyond visit 6 will be assessed clinically. If no recovery is seen, study visits will continue until recovery or discharge. Else, in hospital visits will end with V6; ^1^ Visits may be conducted by phone; ^2^ These assessments are optional, only to be obtained if performed as part of standard of care; ^3^ Only performed if routine blood sampling is indicated. ^4^ Patient reported data that can be collected by phone due to isolation restrictions and follow up visits conducted by phone

|  | Enrol-<br>ment |  |  |  |  |  |  |  |  |  |  |  |
| --- | --- | --- | --- | --- | --- | --- | --- | --- | --- | --- | --- | --- |
| Visit | V1 | V2 | V3 | V4 | V5 | V6 | V7 | V-discharge | V8<sup>1</sup> | V9<sup>1</sup> | V10<sup>1</sup> | V11<sup>1</sup> |
| **Visits will be conducted three times a week until discharge from hospital <sup>5</sup>** | Monday / Wednesday / Friday starting day of enrolment |  |  |  |  |  |  | Day of discharge | 6 weeks after enrolment | 3 months after enrolment | 6 months after enrolment | 12 months after enrolment |
| **Visit Windows** | - | - | - | - | - | - |  | - | +/- 1 week | +/- 2 week | +/- 3 weeks | +/- 3 weeks |
| Informed Consent/Authorization | X |  |  |  |  |  |  |  |  |  |  |  |
| Epidemiological data | X<sup>4</sup> |  |  |  |  |  |  |  |  |  |  |  |
| Demographics | X<sup>4</sup> |  |  |  |  |  |  |  |  |  |  |  |
| Medical History | X<sup>4</sup> |  |  |  |  |  |  |  | X<sup>4</sup> | X<sup>4</sup> | X<sup>4</sup> | X<sup>4</sup> |
| Vital Signs | X | X | X | X | X | X | X | X |  |  |  | X |
| Clinical symptoms | X<sup>4</sup> | X<sup>4</sup> | X<sup>4</sup> | X<sup>4</sup> | X<sup>4</sup> | X<sup>4</sup> | X<sup>4</sup> | X<sup>4</sup> | X<sup>4</sup> | X<sup>4</sup> | X<sup>4</sup> | X<sup>4</sup> |
| Documentation of clinical blood sample results (chemistry, haematology)<sup>2</sup> / Routine blood testing | X | X | X | X | X | X | X | X | X | X | X | X |
| Recording of SARS-CoV-2 PCR testing <sup>2</sup> | X |  |  |  |  | X | X | X |  |  |  |  |
| SARS-CoV-2 Antibody testing <sup>3</sup> | X | X | X | X | X | X | X | X |  |  | X | X |
| Serum cytokines and chemokines, quantitative proteomics, circulating myeloid and lymphoid cells <sup>3</sup> | X | X | X | X | X | X | X | X | X | X | X | X |
| Banked bio-materials <sup>3</sup> | X | X | X | X | X | X | X | X | X | X | X | X |
| Concomitant Medications | X | X | X | X | X | X | X | X | X<sup>4</sup> | X<sup>4</sup> | X<sup>4</sup> | X<sup>4</sup> |
| Documentation of clinical records from patient charts, e.g. parameters of mechanical ventilation, haemodynamics etc. <sup>2</sup> | X | X | X | X | X | X | X | X |  |  |  |  |
| Additional clinical data collection, e.g. Chest X-ray, CT, echocardiography<sup>2</sup> | X | X | X | X | X | X | X | X |  |  |  |  |
| Lung function test, echocardiography |  |  |  |  |  |  |  |  | X | X | X | X |
| Record of any additional hospitalizations or ER visits |  |  |  |  |  |  |  |  | X<sup>4</sup> | X<sup>4</sup> | X<sup>4</sup> | X<sup>4</sup> |
| Clinical severity scores (GCS, SOFA, etc) <sup>2</sup> | X | X | X | X | X | X | X | X |  |  |  |  |
| WHO Clinical Ordinal Scale | X | X | X | X | X | X | X | X | X<sup>4</sup> | X<sup>4</sup> | X<sup>4</sup> | X<sup>4</sup> |
| Quality of Life | X<sup>4</sup> |  |  |  |  |  |  | X<sup>4</sup> | X<sup>4</sup> | X<sup>4</sup> | X<sup>4</sup> | X<sup>4</sup> |

We will also recruit a representative cohort of outpatients with mild symptoms of COVID-19, in whom study visits and biosamplings are only performed at enrolment (V1) and at day 15 (V7) plus follow up, provided that no hospitalization becomes necessary during the course of disease. Telephone interviews will be performed with outpatients in order to assess patient-reported health status between V1 and V7.

#### Serial biosampling, molecular and immunological phenotyping, systematic biobanking

The molecular basis of the widely variable clinical course of COVID-19 is unknown. Many risk factors for severe disease, prognostic biomarkers or correlates of protective immunity remain unclear. Several reports have indicated a central role for improper immune responses in COVID-19 associated lung damage and severe disease [25]. In order to elucidate the immunological basis for different courses of COVID-19 and the development of natural immunity, we have devised a detailed sequential deep phenotyping strategy, to decipher the composition and activation state of cellular and humoral components of the host immune response to SARS-CoV-2, employing high-resolution technologies including single-cell RNA-Sequencing and Cellular Indexing of Transcriptomes and Epitopes by Sequencing (CITE-Seq), T cell receptor- and B cell receptor sequencing, Cytometry by Time-of-Flight (CyTOF), quantitative plasma-proteomics, epigenetic profiling as well as hypothesis-driven investigation and validation of individual biomarkers by conventional immunoassays.

As scientific data on clinical, cellular and molecular aspects of COVID-19 are rapidly evolving, we will preserve high-quality biosamples, including cryopreserved immune cells, through a large-scale biobanking effort. An overview of the sample-flow is shown in figure 1. Blood samples are collected three times per week in hospitalized patients, alongside blood drawing for routine laboratory analysis. Biobanking is organized in close cooperation with central biobank of Charité/BIH (ZeBanC) according to standard operational procedures (SOPs) and under certified conditions.

**Figure 1:**
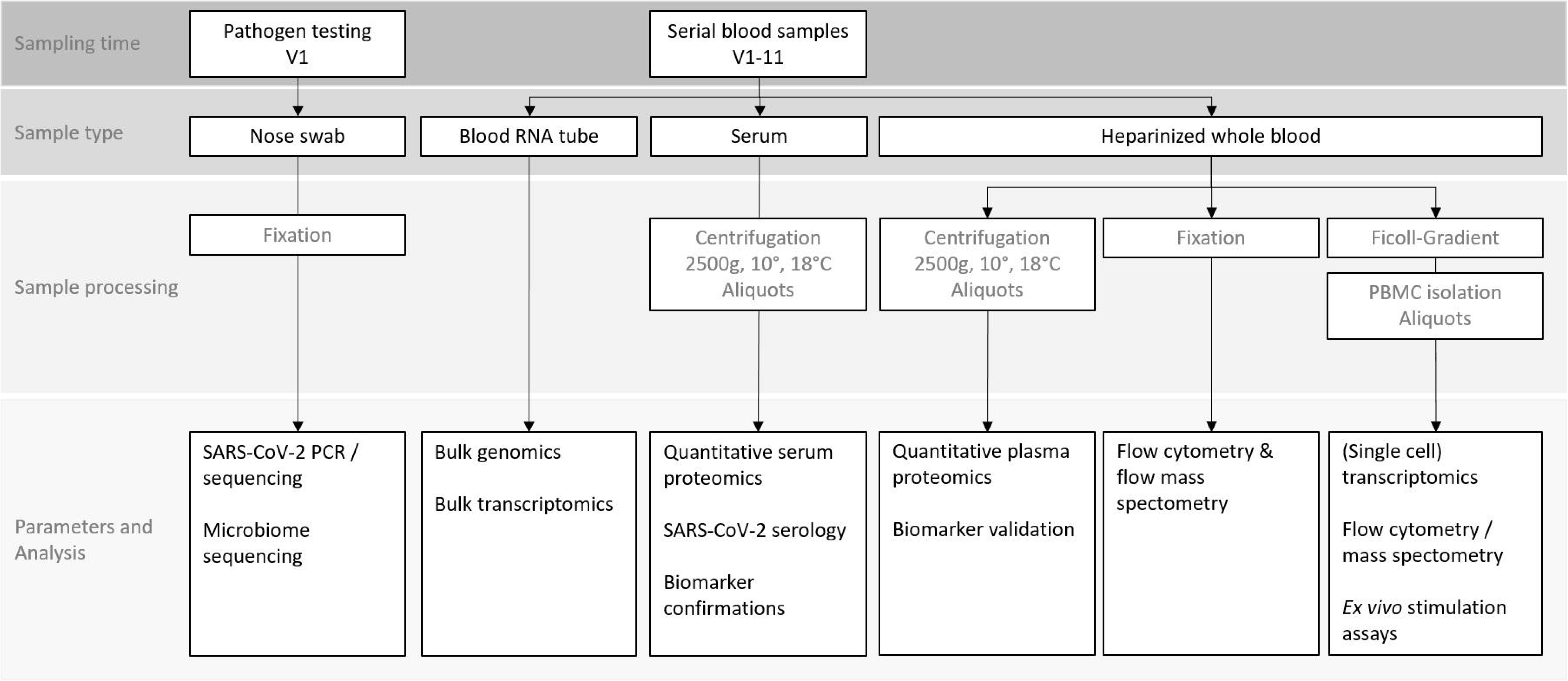
Depiction of serial biosampling and analysis performed during study conduction; Pathogen testing will be performed only at first study visit (V1), while serial blood sampling will be performed repetitively at every study visit (V1-V13).

To ensure maximal safety for medical staff involved in this clinical study, SOPs for clinical sample handling are defined. Personal safety equipment for laboratory staff includes tightly closed protective gown, face shield or protective glasses, surgical face mask and protective gloves. All blood samples are handled under a bio-safety level (BSL) 2 hoods and primary tubes are discharged. Analyses of infectious specimen will be handled according to current regulation under BSL2 safety conditions.

#### Post-mortem analysis

Post mortem analysis under strict safety considerations will be considered for patients succumbing to COVID-19 in this study. A detailed SOP has been devised at our institution. Autopsy will provide in-depth insights into viral tropism and organ pathologies of severe disease. Additional analysis of infected tissue using spectral confocal microscopy will provide insights into the pathology of fatal COVID-19. Samples will be stored for future analysis.

### 2.5 Data management and -storage

Clinical and laboratory data will be collected from patient charts, through patient interviews or from examinations and submitted online to a purpose-built eCRF using the electronic data capture system SecuTrial®. All data will be pseudonymized using a 6-digit alphanumerical patient code and the online records kept will not include any information that allows patient identification. An enrolment log list will be used to match patient codes in the database to individual patients in order to record clinical outcomes and supply any missing data points. The enrolment log and study data will be kept separately. Access to the database is protected by username and password. The electronic database set up in this study is suitable for the use in interventional trials and can therefore rapidly be modified for future clinical testing of therapeutic agents.

### 2.6 Quality assurance and control measures

A detailed data dictionary will define data to be collected on eCRF. Quality checks will be built into the data management system, including the possibility to generate queries, in order to ensure standardization and validity of the data collected. Any information that is not available to the investigators will be considered as missing. No assumptions will be made for missing data. Data monitoring of a randomly selected subset of collected data will be performed in the course of the study. Discussion of data collection techniques throughout the course of the study with investigators will ensure data quality and allow for adjustments.

### 2.7 Data storing, access and –sharing

Access to samples for additional analyses, open access and data sharing via the Global Research Collaboration for Infectious Disease Preparedness (GloPID-R) will be governed by a research committee comprising the clinical lead investigators and scientific investigators for this study. It is a fundamental principle of this study that all contributors and researchers who have access to samples commit to unrestricted data sharing in order to provide timely insights into this new disease. In addition, all clinical investigators contributing to this research efforts will be given full recognition for their efforts and the opportunity to access data and samples. Data management adheres to the FAIR data principles, stating that all data collected shall be findable, accessible, interoperable and re-usable [26]. Samples collected will be used for the purpose of this study as stated in the protocol and consented for future use.

### 2.8 Ethics and registration

This study is conducted in compliance with the principles laid down in the 1964 Declaration of Helsinki and its later amendments. Where applicable, the principles of Good Clinical Practice (International Council for Harmonization, ICH 1996) and other applicable regulations and guidelines will be used to guide procedures and considerations. The study was reviewed and approved by the Charité Ethics Committee (EA2/066/20). All patients enrolled will give written informed consent in person, or by a legal guardian. Pa-COVID-19 is currently being registered at clinicaltrials.gov.

Medical management of participants in this study must never be compromised by study procedures. At all times, priority will be given to samples required for medical management and day-to-day care of patients. Study blood samples will be taken together with routine blood sampling for clinical monitoring in order to minimize the individual risk connected with phlebotomy or manipulation of intravascular catheters. There is no direct benefit for patients participating in the study, but results from the study might improve clinical care in future patients and benefit public health.

Specific ethical consideration includes *the recruitment of critically ill patients* who might not be able to consent, *the strong moral obligation to participate* in a public health emergency*, keeping the balance between public health and research*, and the *potential risk for research staff* involved in this study.

The enrolment of patients, who are unable to consent is a common challenge in critical care, acute research and minors. The process follows a clear legal framework with consent being obtained from legal guardians as soon as possible. In addition, all efforts will be made to obtain informed consent from patients retrospectively after recovery. For patients refusing retrospective informed consent, all data and biosamples will be deleted. For patients succumbing to COVID-19 before informed consent can be obtained, all data will be anonymized.

The patient information clearly outlines potential benefits and limitations. The autonomy and wellbeing of the individual patient and the medically indicated procedures and treatments are always paramount. All staff is trained in infection control measures and have ready access to appropriate personal protective equipment. Dedicated research staff will be available to support the study activities.

## 3. Conclusion

This study protocol of the observational prospective cohort study Pa-COVID-19 that has recently started recruitment at Charité - Universitätsmedizin Berlin may serve as a blueprint for data and biosample collection in patients with COVID-19 in Germany. The protocol combines a comprehensive clinical and translational approach with tools for implementation in different clinical settings. It is scalable to local resources, and by design, this platform allows for rapid initiation of interventional trials, which are urgently needed to investigate effective treatments for COVID-19.

**Supplemental material:** Data collection elements for scalable data collection. Data elements are categorized to allow scalability to local research resources. Categories are: core data elements (1), recommended data elements (2) and supplemental data elements (3).

## Data Availability

the study protocol shall serve as a blueprint for studies conducted on patients infected with SARS-CoV-2 in Germany.

https://studycenter.charite.de/covid_19/

